# SARS-CoV-2 epidemic in the South American Southern cone: can combined immunity from vaccination and infection prevent the spread of Gamma and Lambda variants while easing restrictions?

**DOI:** 10.1101/2021.09.16.21263701

**Authors:** Marcelo Fiori, Gonzalo Bello, Nicolás Wschebor, Federico Lecumberry, Andrés Ferragut, Ernesto Mordecki

## Abstract

All South American countries from the Southern cone (Argentina, Brazil, Chile, Paraguay and Uruguay) experienced severe COVID-19 epidemic waves during early 2021 driven by the expansion of variants Gamma and Lambda, however, there was an improvement in different epidemic indicators since June 2021. To investigate the impact of national vaccination programs and natural infection on viral transmission in those South American countries, we analyzed the coupling between population mobility and the viral effective reproduction number *R*_*t*_. Our analyses reveal that population mobility was highly correlated with viral *R*_*t*_ from January to May 2021 in all countries analyzed; but a clear decoupling occurred since May-June 2021, when the rate of viral spread started to be lower than expected from the levels of social interactions. These findings support that populations from the South American Southern cone probably achieved the conditional herd immunity threshold to contain the spread of regional SARS-CoV-2 variants.

## 1 Introduction

Countries from the South America Southern cone experienced large COVID-19 epidemic waves during the first months of 2021 driven by the lack of stringent mitigation measures along with the emergence and regional spread of the Variant of Concern (VOC) Gamma and the Variant of Interest (VOI) Lambda [1]. The VOC Gamma was the predominant viral variant in Brazil, Paraguay and Uruguay; while both Gamma and Lambda circulated at similar prevalence in Argentina and Chile [2, 3, 4, 5]. Changes in different epidemic indicators from mid-June to end of August, including declining numbers of new SARS-CoV-2 cases and deaths and viral effective reproduction number *R*_*t*_ below one, support a relative control of the COVID-19 epidemic in all five countries [1]. The drivers of such epidemic control remained unclear as SARS-CoV-2 transmission could be influenced by several factors including extent of non-pharmaceutical interventions (NPIs), level of social distancing, adherence to self-care measures, transmissibility of circulating viral variants and the proportion of susceptible host [6].

Several studies demonstrate that during the pre-vaccination phase and in a context of large community transmission of the virus, when other factors as contact tracing strategies are not effective, changes in population mobility could be predictive of changes in epidemic trends and viral *R*_*t*_ [7, 8, 9, 10, 11, 12, 13]. In those settings, decoupling between population mobility and viral transmissions could be used as a surrogate marker of herd immunity achieved either through high vaccination and/or natural infection rates. Data from countries with advanced vaccination like Israel and the United Kingdom support this notion as in a certain time SARS-CoV-2 incidence display sustained declines despite easing of lockdown restrictions, discontinuation of face mask use in open spaces and increase in population mobility [14, 15]

In the present article, we estimate the coupling between population mobility and the *R*_*t*_ of SARS-CoV-2 in the five South American countries from the Southern cone. Our analyses support that mobility data was highly correlated with the viral *R*_*t*_ in all South American countries analyzed between January and May, 2021; however, a clear decoupling between population mobility and viral transmissions was evident since May-June 2021. The mean estimated threshold of immune individuals (fully vaccinated pondered by vaccine effectiveness plus natural infected) necessary to produce such decoupling varies along the five countries from 29% to 45% and a discussion trying to understand these differences is provided. These findings also support the relevance of vaccination-induced herd immunity in South American countries with widespread use of the inactivated vaccine Coronavac.

## 2 Results

To analyze the potential correlation between social mobility and the spread of the SARS-CoV-2, we estimate the viral effective reproduction number *R*_*t*_ in every country based on mobility information provided by Google [16] during a time period of high viral transmission (see subsection 4.2). The resulting estimator, denoted as 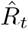, was then correlated with the observed *R*_*t*_ estimated from the incidence data available in the Our World in Data (OWID) data base [1].

The correlation between 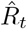 and *R*_*t*_ provides a measure of the value of social mobility as a predictor of viral transmissions in each country, while the ratio 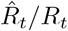 provides a measure of the coupling between both indicators. In all five South American countries analyzed (Argentina, Brazil, Chile, Paraguay and Uruguay) we observed that during the first months of 2021, the estimated 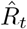 was highly correlated (*ρ*^2^ between 0.83 y 0.94) with the observed *R*_*t*_ about 1-weeks later and the ratio 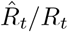 was close to one (0.90-1.10) during the pre-vaccination and initial vaccination phases (Figure 1). We observed a high correlation between both estimators not only during the estimation period, but also during the beginning of the vaccination roll-out. These findings confirm that population mobility was a relevant driver of viral transmissions during early 2021 in all South American countries analyzed and revealed that, under a context of high community transmission, researchers can use the observed population mobility at a given time to infer the viral transmission dynamics without the typical lag of the observed *R*_*t*_.

**Figure 1:**
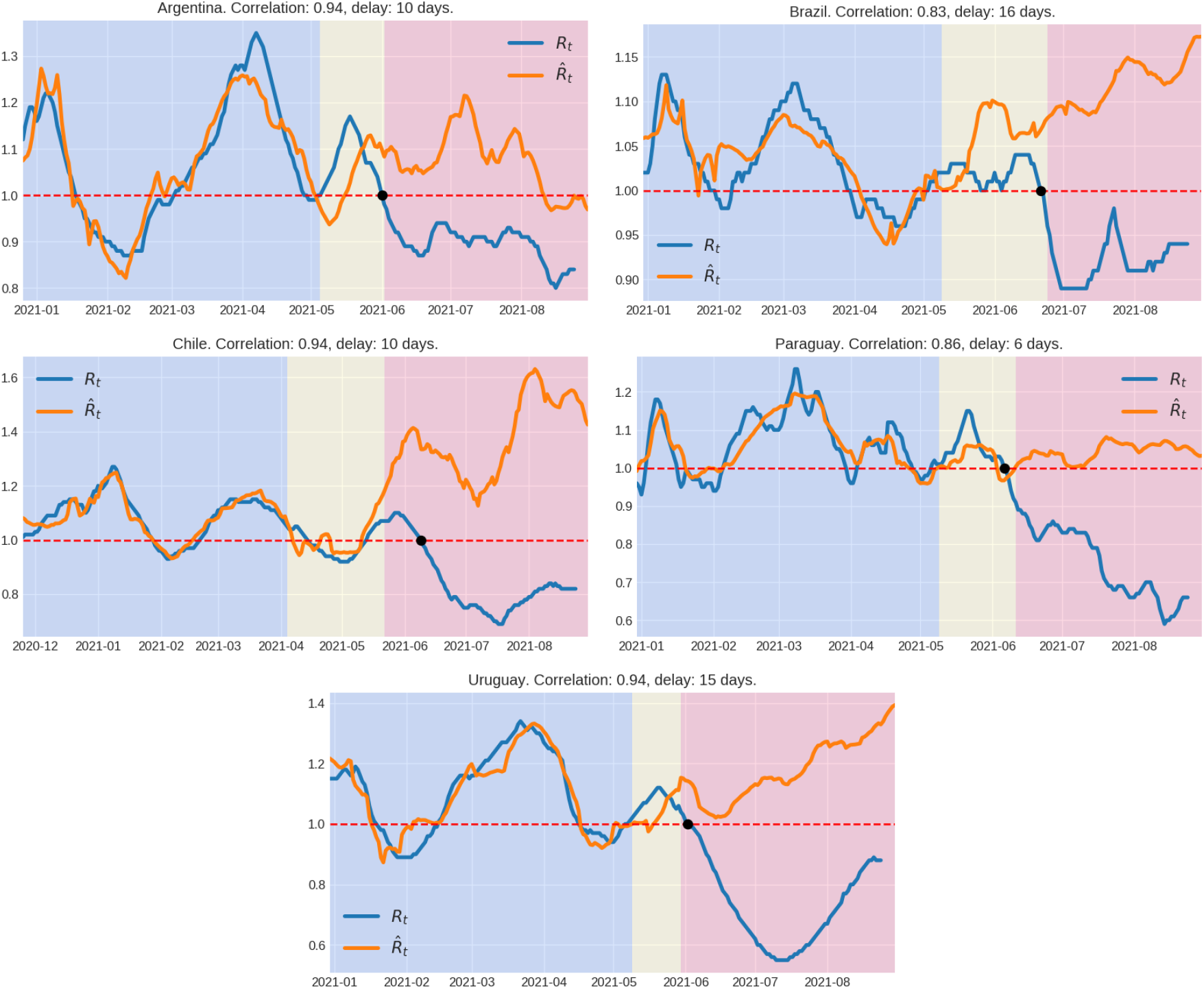
Viral effective reproduction number *R*_*t*_ and its estimation 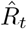 using mobility information. Background colors indicate the following time periods: in blue, the time period used to fit the linear model (see Section 4.2), in yellow, the period after the fitting, but before the decoupling time, and in red after the decoupling point. The black dot corresponds to the last time the reproductive number was above one. The correlation corresponds to the period used to fit the model. The delay indicated is the time-shift between the mobility time series and *R*_*t*_ in order to maximize the correlation in the linear regression.

When we extended the estimation of the 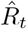 during the vaccination roll-out period (with the same computed initial parameters), we observed a clear increase of the ratio 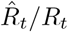 in all South American countries analyzed since late May and early June 2021, indicating that at a certain time the rate of spread of the virus started to be lower than expected from the levels of social interactions (Figure 1). We interpret such decoupling between population mobility and viral spread as a surrogate marker of conditional herd immunity, i.e. the achieved herd immunity conditioned to the social distancing policies and the circulating viral variants in each country. In order to test our method, we conducted a similar analysis in Israel, the first country to attain conditional vaccine-induced herd immunity. Our findings confirm that after a period of clear coupling between population mobility and viral transmission, a decisive increase of the ratio 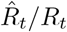 was also observed at a certain time during vaccination roll-out in Israel (Figure A.1). The decoupling time, defined as the moment when the ratio 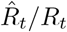 finally overcomes (i.e. the last time it crosses) the value 1.10, preceded the last peak of weekly reported cases and roughly coincides with the last day when the *R*_*t*_ = 1 in each country (Figure 1), indicating that the decoupling time was an early indicator of epidemic control.

The proportion of immunized population at the decoupling time could give us an idea of the conditional herd immunity threshold (HIT). In order to estimate the proportion of immune individuals around the decoupling time, we summed the estimated number of vaccine-immunized and natural-immunized individuals. The proportion of vaccine-immunized individuals was estimated from the number of fully vaccinated individuals adjusted by the estimated vaccine effectiveness (VE) in South America [17, 18], see also [19]. The number of infected people that acquired immunity through previous infection (cumulative infection) was estimated from the cumulative number of deaths assuming a constant (age adjusted) infection fatality rate (IFR) for each country (see subsection 4.1 and Table 1). The mean estimated HIT at the decoupling time varies along the countries from 29% in Argentina to 33% in Uruguay, 36% in Paraguay, 43% in Chile and 45% in Brazil, although confidence intervals were very large due to uncertainties in the IFR estimates (Table 1 and Figure 2). The HIT was reached by different proportions of natural infections and vaccination (Table 1). The estimated proportion of individuals that acquired immunity through vaccination (taking into account the VE) at the decoupling time was relatively high in Chile (29%) and Uruguay (24%), but very low in Brazil (9%), Argentina (5%) and Paraguay (1%). The estimated HIT in countries with widespread use of the inactivated vaccine Coronavac like Chile (43%) and Uruguay (33%) was similar to that estimated in Israel (42%) that only used the BNT162b2 (mRNA-based) vaccine (Figure A.2).

**Table 1:** IFR: infection fatality rate; VIN: percentage of virus inactivated vaccines; ADV: percentage of adenovirus vaccines; RNA: percentage of RNA vaccines [20, 21, 22, 23, 24, 25]; Dec-T: decoupling time; % Nat-Inf: percentage of population naturally infected at Dec-T; % Vac: percentage of the population fully vaccinated at Dec-T; HIT (herd immunity threshold): percentage of immunized population due to vaccines and natural infections at Dec-T. The vaccine effectiveness (VE) against SARS-CoV-2 infections was adjusted to 66% for VIN, 73% for ADV and 93% for RNA [17, 18].

| Country | IFR | (VIN, ADV, RNA) | Dec-T | % Nat-Inf | % Vac | HIT |
| --- | --- | --- | --- | --- | --- | --- |
| Argentina | 0.67 (0.36-1.30) | (31.1, 64.7, 04.2) | Jun. 02 | 26 (13-48) | 06 | 29 (17-52) |
| Brazil | 0.59 (0.32-1.17) | (34.4, 48.1, 17.5) | Jun. 23 | 40 (20-74) | 11 | 45 (25-79) |
| Chile | 0.73 (0.40-1.43) | (71.1, 06.9, 22.0) | May 22 | 20 (10-37) | 40 | 43 (34-60) |
| Paraguay | 0.41 (0.23-0.83) | (11.6, 26.6, 61.8) | Jun. 11 | 35 (18-64) | 02 | 36 (19-64) |
| Uruguay | 0.90 (0.49-1.56) | (59.8, 01.6, 38.6) | May 29 | 13 (8-24) | 29 | 33 (27-44) |
| Israel | 0.65 (0.35-1.27) | (0,0,100) | Feb. 28 | 10 (5-19) | 39 | 42 (37-51) |

**Figure 2:**
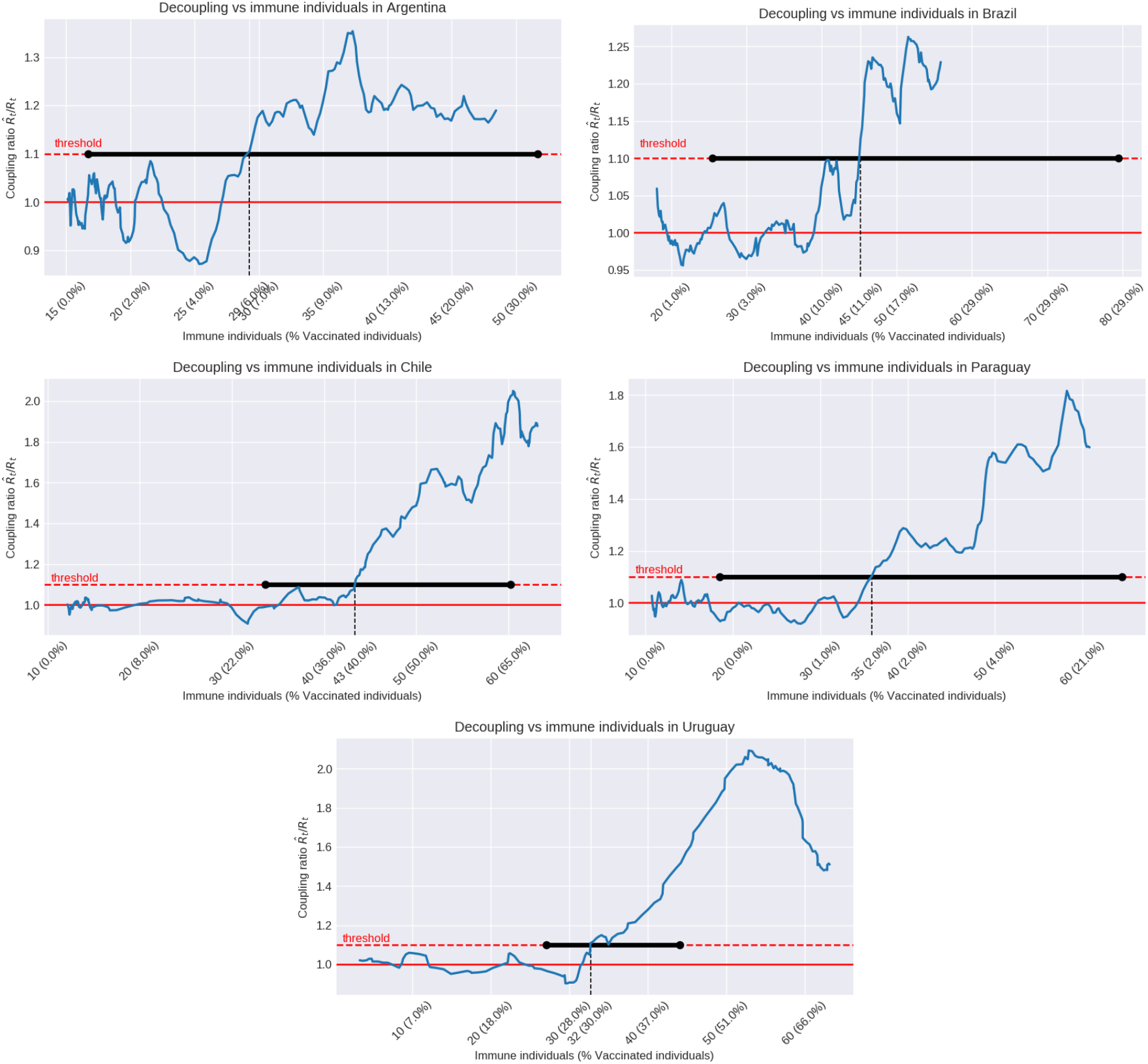
Coupling ratio 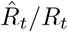 plotted with respect to the percentage of immune population. During the first months of 2021 the coupling ratio varies around 1, which corresponds to the periods where the *R*_*t*_ and 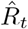 are in concordance in Figure 1. Immune population includes immunity achieved by vaccination (taking into account its effectiveness), and natural infection (see subsection 4.3). The percentage of people fully vaccinated is described as well. The coupling ratio crosses the threshold (decoupling point) at percentages of immune population that varies along the five countries from 29% in Argentina to 33% in Uruguay, 37% in Paraguay, 43% in Chile and 45% in Brazil. Confidence intervals are shown in horizontal black lines. They inherit the large uncertainty in the IFR estimation (see Table 1).

## 3 Discussion

All countries from the South America Southern cone (Argentina, Brazil, Chile, Paraguay and Uruguay) witnessed pronounced increases in daily SARS-CoV-2 cases and deaths during the firsts months of 2021 and a clear drop in relevant epidemic metrics (cases, deaths and *R*_*t*_) from mid-2021 [1]. This study demonstrates that such epidemic control was preceded by a clear decoupling of viral transmissions from population mobility, consistent with the notion that those South American countries probably attained the HIT against SARS-CoV-2 variants Gamma and Lambda prevalent in the region, given some level of social dis tancing restrictions.

At the start of the pandemic, thresholds of 60-70% were given as estimates of herd immunity for SARS-CoV-2 [26]. Despite confidence intervals of HIT estimates were very large, mostly due to uncertainties in the IFR estimates, our analyses support that the conditional HIT for SARS-CoV in South America would be lower than 50%, ranging from 29% in Argentina to 45% in Brazil. Moreover, observe that these confidence intervals have a common range of (34, 44) = 39±5. A recent modeling study conducted in Stockholm, Sweden, also supports that this country reached the HIT against the original and Alpha variants of SARS-CoV-2 at 23% and 33% of seroprevalence, respectively [27]. The authors conclude that HIT for SARS-CoV-2, given limited social distancing restrictions, could be lower than initially estimated and that phenomena could be explained by population heterogeneity. By fitting epidemiological models that allow for heterogeneity in susceptibility or exposure to SARS-CoV-2 and given a basic reproduction number *R*_0_ between 2.5 and 3, a recent study estimates that the HIT declines from over 60% to less than 10% as the coefficient of variation increases [28]. Another study estimate that in an age-structured community with mixing rates fitted to social activity, the HIT can be 43% if *R*_0_ is 2.5 [29].

Our findings also support that the conditional HIT for SARS-CoV-2 in South America was attained through both natural and vaccinal immunity, with different relative proportions across countries. The extremely low proportion of vaccine-immune individuals in Paraguay (1%), Argentina (5%) and Brazil (9%) at decoupling time, suggest that conditional herd immunity in those countries was mostly attained by natural infections. Few studies estimated the proportion of infected individuals in South America after the large Gamma and Lambda epidemics in 2021, but some evidence from seroprevalence data support our estimations. A randomized study conducted in Paraguay between March to June 2021 gave a seroprevalence of 23.1% in Asunci’
son and of 26.9% in the central region of the country [30] and a recent seroprevalence survey among adult individuals living in the largest Brazilian city of Sao Paulo also estimate a high proportion (45%: 39-51%) of individuals infected by SARS-CoV-2 [31].

At the other extreme, the relative proportion of vaccinal immunity at decoupling was highest in Chile (29%) and Uruguay (24%). CoronaVac accounted for most of vaccinations in Chile (75%) [32] and Uruguay (66%) [24] and the high incidence of SARS-CoV-2 in those countries during first months of vaccination roll-out raise concerns about the effectiveness of inactivated virus vaccines to control SARS-CoV-2 transmissions. Our results support that the widespread use of inactivated virus vaccines contributed to containing the spread of SARS-CoV-2 in Chile and Uruguay, despite abundant circulation of VOCs/VOIs and weak mitigation measures. Remarkably, the HIT at decoupling point in Chile (43%) and Uruguay (33%) was similar to the one estimated for Israel (42%), that mostly controlled the virus expansion through vaccination with BNT162b2. These findings are consistent with recent studies of vaccine effectiveness (VE) in Chile [17], Brazil [18] and Bahrain [33] that conclude that immunization with inactivated vaccines (CoronaVac and Sinopharm) was an effective strategy at mitigating the risk for transmissions of SARS-CoV-2 VOCs, although the performance of BNT162b2 and adenovirus-based vaccines was superior.

The mean estimated HIT varied across South American countries and several factors may explain such variability. HIT will move upwards when more transmissible SARS-CoV-2 variants circulates in a population, but differences in the circulating SARS-CoV-2 variants do not explain variations among South American countries. Differences in the mean HIT were observed between countries where Gamma was the most prevalent variant like Brazil (45%), Paraguay (36%) and Uruguay (33%), and also between countries where Gamma and Lambda co-circulated at high prevalence like Chile (43%) and Argentina (29%) [2, 3, 4, 5]. Differences in vaccine platforms deployed in each country might also modulate the HIT at the decoupling time. Although we corrected the proportion of immune individuals according to the estimated VE and the proportion of each vaccine, we only considered immunity associated with fully vaccinated individuals. Previous studies, however, demonstrate some level of reduction of SARS-CoV-2 transmission after one dose of mRNA-based (46-58%), adenovirus-based (35%) and inactivated virus (16%) vaccines [17, 18, 34, 35]. Thus, we should expect that countries that used a higher proportion of mRNA-based and/or adenovirus-based vaccines like Argentina (69%) reached herd immunity at apparent lower thresholds that those that mostly used inactivated virus vaccines. Moreover, it should be stressed that Argentina had a very large proportion of individuals with a single dose at the decoupling point when compared to other countries in the region where second doses were administrated in a shorter period after first dose [1]. Notably, although Brazil also used an overall high proportion of mRNA-based and/or adenovirus-based vaccines (66%), most vaccinations during first months were of inactivated vaccines [18].

Reduction of SARS-CoV-2 transmission will also depend on the vaccination strategy (who is vaccinated and when). Vaccinations programs usually begin by elderly people and go on by gradually protecting the younger population [36]. Simulation studies indicate that prioritize vaccinating of high-risk groups will minimize the number of COVID-19-related hospitalizations and deaths in the short term, but vaccination of main transmission drivers (i.e. highly mobile working age groups) would be more effective at reducing the spread of the SARS-CoV-2 [37, 38]. Given enough vaccine supplies, vaccinating the adult population uniformly at random would thus be ideal to both prevent death and severe illness in high risk groups and to curb SARS-CoV-2 transmissions in the whole population. Uruguay developed an interesting vaccination strategy that prioritized vaccination of elderly populations (≥70 years of age) with the BNT162b2 vaccine while highly mobile working age groups were simultaneously vaccinated with CoronaVac. This more homogeneous vaccination strategy across different age groups in Uruguay might partially explain the relative low HIT observed in this country. This may be related to the fact that, the decoupling effect due to vaccinations programs that we observe between mobility and the reproductive number is reached more abruptly than what could be expected from SIR-like models where all the population is treated homogeneously.

Our results support that proportion of immune population in South American populations attained a threshold enough to decoupling people mobility and viral dissemination and those countries could thus implement progressive relaxing of mitigation measures with relative safety. Such apparent herd immunity, however, was attained while maintaining moderate mitigation measures (social distancing, school closed, mask-wearing and other self-care behaviors). None of the countries analyzed have returned to the prepandemic levels of activity and it is unclear if current population immunity will halt the viral spread after removal of all mitigation measures. Long-term herd immunity could be also challenged by waning immunity and dissemination of more infectious SARS-CoV-2 variants [39]. Waning neutralizing antibodies might progressively reduce the population immunity level to below the critical HIT, while local evolution and/or introduction of SARS-CoV-2 variants that are more transmissible than those previous circulating will move the HIT upwards.

Both factors seems to have shaped the third epidemic wave in Israel [40, 41, 42, 43] Our study supports that after a transient period of decoupling in Israel, population mobility and viral transmissions were coupled again as Delta variant spread in both unvaccinated and fully-vaccinated individuals. It is unclear if the same phenomena could be observed in South America after introduction of Delta variant. First, herd immunity through natural infection seems to be less susceptible to waning immunity than by vaccination [44, 45, 46] and South American countries with a high natural immunity wall might be better prepared to limit the expansion of Delta variant than those with a large vaccine immunity wall. Second, hybrid immunity (natural infection plus vaccination) might provide longer lasting and stronger protection against infection than vaccine-induced immunity [47] and a high proportion of partial or fully vaccinated individuals in South America may be currently in this condition. Third, some South American countries like Chile, Uruguay and Brazil already started or approved the administration of a vaccine booster.

Our study has some important limitations: (i) difficulty to estimate precisely the IFR and consequently to have a precise estimate of the cumulative number of naturally infected people at decoupling point in each country; (ii) sub-reporting of SARS-CoV-2 deaths might underestimate the cumulative number of infections and thus the HIT; (iii) the assumption that partially vaccinated people did not greatly contribute to reduce viral transmissions might have also underestimate the number of vaccine-immune individuals and the actual HIT; (iv) on the other hand, although we assumed some overlap between vaccinal immunity and natural immunity, the precise fraction of fully vaccinated individuals that were previously infected is unknown. Because of these limitations, the precise HIT estimated here should be interpreted with caution and should not be considered as general reference values for other countries.

In summary, our study supports that populations from the South American Southern cone probably achieved the conditional HIT to contain the further spread of SARS-CoV-2 variants Gamma and Lambda at around mid-2021. Presumed herd immunity was probably mostly attained by natural infection in Argentina, Brazil and Paraguay, and by a mixture of natural infections and vaccination in Chile and Uruguay. The widespread used of the Coronavac inactive viral vaccine in South America was not only effective to prevent the severe forms of COVID-19 disease but also has the potential to contain the community spread of highly transmissible SARS-CoV-2 regional variants. Inactivated SARS-CoV-2 vaccines, combined with other vaccines and mitigation measures, may thus represent a relevant tool to control the COVID-19 pandemic especially under the severe limitation of vaccine supplies faced by many countries around the world. Our findings stress that the herd immunity status might be rapidly lost if vaccine-induce neutralizing antibodies decrease over time and more transmissible SARS-CoV-2 variants are either introduced from abroad or evolved locally.

## Data Availability

All the data and code used in this paper are publicly available, and linked along the text.

https://github.com/marfiori/covid19-decoupling

## Acknowledgements

We are indebted to P. Bermolen, M. Cerminara, M. I. Fariello, F. Paganini, R. Radi, M. Reiris, A. Rojas de Arias and C. E. Schaerer, for useful discussion and support. Any errors are the responsibility of the authors.

## 4 Methods

### 4.1 Data and code availability

The SARS-CoV-2 incidence data, viral effective reproduction number *R*_*t*_ (also indicated as reproduction rate), confirmed deaths, vaccinated people, and other epidemiological indicators were retrieved from Our World in Data (OWID) [1]. Mobility index was estimated from the six indicators categories (retail and recreation, grocery and pharmacy, parks, transit stations, workplaces, and residential) provided by Google COVID-19 Community Mobility Reports [48]. For the sake of reproducible research, the code used to obtain all the results and figures is available at https://github.com/marfiori/covid19-decoupling.

### 4.2 Estimation of the viral effective reproduction number and decoupling time

As the correlations between the six different possible regressors are large, we construct indices that are more robust along time and different countries, to avoid overfitting. In order to do this, we choose for each country the three categories that give the best fit among all possible combinations. Although the categories may vary, the obtained fit quality is relatively robust over different time intervals. The six mobility time series were smoothed by averaging over a 14 days sliding window.

For each country, we selected a time period consisting of 75 days before the start of the vaccination campaign, and 55 days after, ending up with a 130-days period to carry out the estimation. Given a set of three mobility categories, we fitted a linear regression model to the viral effective reproduction number *R*_*t*_, lagged a certain time period. This time shift was chosen as the lag that maximizes the correlation of the regression. This procedure was repeated for each combination of three categories among the six mobility measures provided by Google, and the combination achieving the best regression result was kept. It should be noted that, since the six categories are highly correlated, other combinations of three categories achieve similar fitting results, and therefore the chosen categories are not necessarily informative by themselves.

Using the coefficients obtained in this 130-days period, and rest of the mobility time series, we computed the predicted viral reproduction number 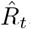. The procedure was tested using periods of different lengths for the estimation, and the results in the HIT are robust along the different experiments.

When population mobility and viral transmission are coupled, the coupling ratio 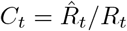 oscillates around one (0.90-1.10). Departing from a certain moment, the 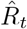 becomes much higher than the *R*_*t*_, revealing the decoupling between population mobility and viral spread resulted. We defined the **decoupling time** *D*_*t*_ as the moment when the coupling ratio 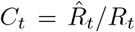 definitely exceeds the value 1.10, i.e. the last crossing over 1.10.

### 4.3 Estimation of the IFR and immune population

As it is well known, the estimation of the infection fatality rate has been a hard task during all the pandemic. The cryptic circulation of the virus (due to asymptomatic infections) and different variants made that in fact this quantity varies along time and populations. Here we took into account the most relevant variable to compute it, that is the age structure of the population. We then took IFR by age taken from [49] and adjusted to the population pyramid of each of the considered countries [50]. Confidence intervals were calculated by considering the (very large) confidence intervals available from [49] and estimating the interval for the whole population as the weighted average of the positions for the maximum or minimum of the age-classes intervals. Only one exception was introduced: in the Uruguayan case, the confidence interval can be reduced because the IFR must be smaller than the Case Fatality Rate (CFR). Imposing this constraint the maximum possible value in the Uruguayan case is reduced (we obtained the CFR corresponding to July 31 from [1]) the other countries being unaffected. This IFR estimation was confirmed using an alternative methodology for the case of Uruguay, following [51], which led to similar results, but with slightly larger confidence intervals.

The percentage of immune population was computed considering the immunity achieved by vaccination (including its effectiveness), and natural infection. However, many people who gained immunity by natural infection, might have gotten vaccinated as well. In order to avoid the over estimation resulting from counting twice those subjects, we subtracted the intersection of these fractions, under the assumption that they are independent. Observe that this assumptions gives us a lower bound on the estimation of immune population.

For a given country, let us denote by *FV* the proportion of fully vaccinated people, by *NI* the proportion of people with immunity by natural infection, and by *V E* the vaccine effectiveness of the country, computed by combining the effectiveness of each vaccine type (VIN, ADV, RNA) using the proportion of vaccines used in the country (see Table 1). We assumed a perfect immunization due to natural infection. That is, we neglected in the present analysis the number of re-infections. Furthermore, let us denote by *IM* the estimation of the proportion of immunized population. Then, the computation described above is as follows:

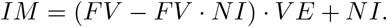

Here the product *FV* · *NI* accounts for the intersection of the populations, which is subtracted from the vaccinated population before the effectiveness factor is applied. As described through the text, the proportion of people with immunity by natural infection is inferred from the confirmed deaths, using the estimated *IFR*.

Observe that due to the vaccine effectiveness, the percentage of fully vaccinated people may by greater than the percentage of immunized population.

## A Supplementary Material

In figures A.1 and A.2 we provide the same analysis shown in figures 1 and 2 in the case of Israel, respectively.

**Figure A.1:**
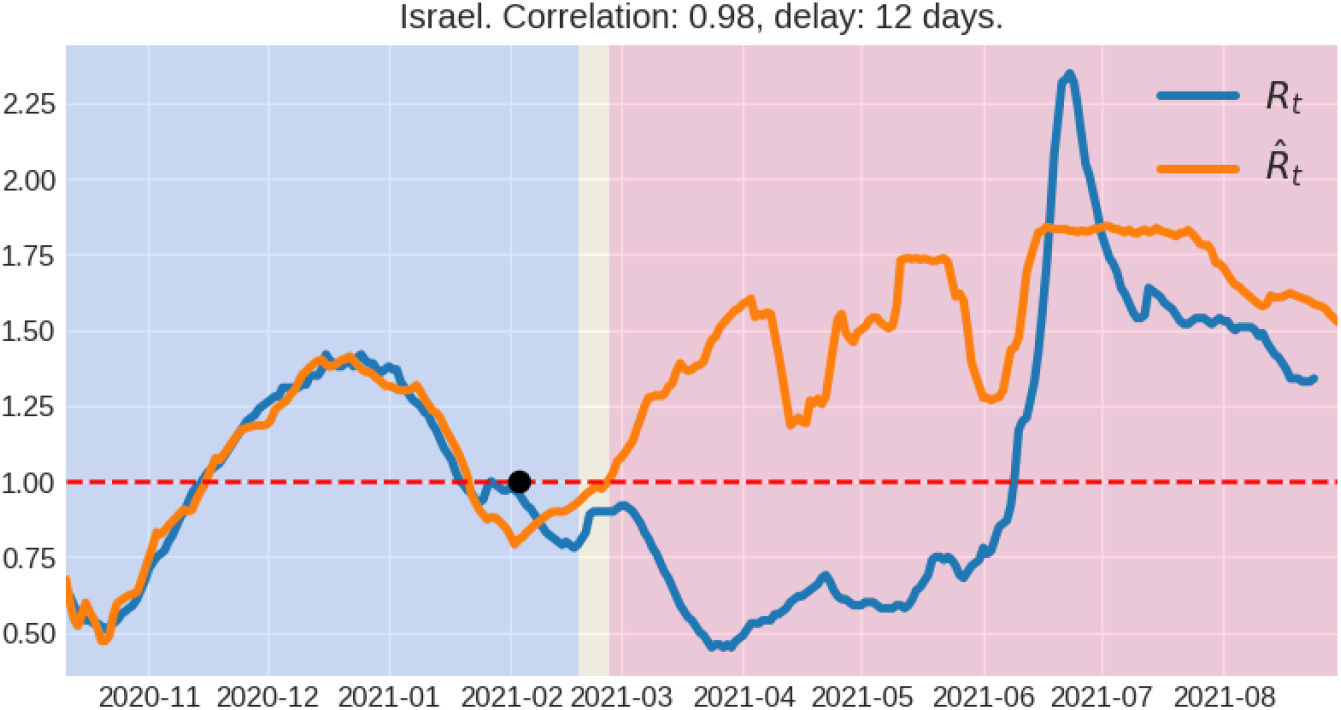
Viral effective reproduction number *R*_*t*_ and its estimation 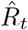 using mobility information. Background colors indicate the following time periods: in blue, the time period used to fit the linear model (see Section 4.2), in yellow, the period after the fitting, but before the decoupling point, and in red after the decoupling point. The black dot corresponds to the last time the reproductive number was above one. The correlation corresponds to the period used to fit the model. The delay indicated is the time-shift between the mobility time series and *R*_*t*_ in order to maximize the correlation in the linear regression.

**Figure A.2:**
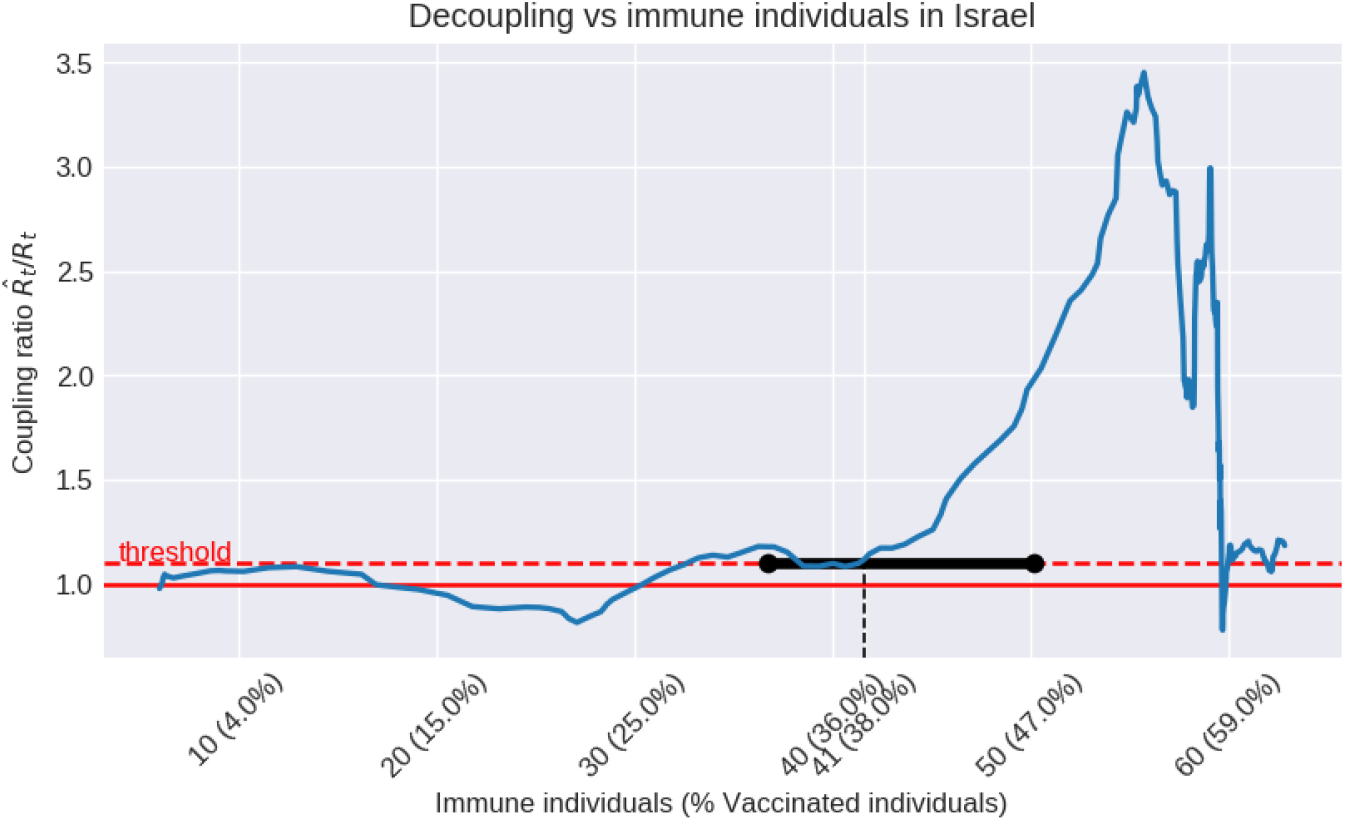
Coupling ratio 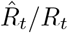 plotted with respect to the percentage of immune population. During the first months of 2021 the coupling ratio varies around 1, which corresponds to the periods where the *R*_*t*_ and 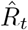 are in concordance in Figure A.1. Immune population includes immunity achieved by vaccination (taking into account its effectiveness), and natural infection (see subsection 4.3). The percentage of people fully vaccinated is described as well.

